# SIREN protocol: Impact of detectable anti-SARS-CoV-2 on the subsequent incidence of COVID-19 in 100,000 healthcare workers: do antibody positive healthcare workers have less reinfection than antibody negative healthcare workers?

**DOI:** 10.1101/2020.12.15.20247981

**Authors:** S. Wallace, V. Hall, A. Charlett, P.D. Kirwan, M.J. Cole, M. Shrotri, S. Rokadiya, B. Oguti, A. Vusirikala, M. Zambon, T. Brooks, M. Ramsay, C.S. Brown, M.A. Chand, S. Hopkins

## Abstract

**Background:** The overall risk of reinfection in individuals who have previously had COVID-19 is unknown. To determine if prior SARS-CoV-2 infection (as determined by at least one positive commercial antibody test performed in a laboratory) in healthcare workers confers future immunity to reinfection, we are undertaking a large-scale prospective longitudinal cohort study of healthcare staff across the United Kingdom.

**Methods:** Population and Setting: staff members of healthcare organisations working in hospitals in the UK

At recruitment, participants will have their serum tested for anti-SARS-CoV-2 at baseline and using these results will be initially allocated to either antibody positive or antibody negative cohorts. Participants will undergo antibody and viral RNA testing at 1-4 weekly intervals throughout the study period, and based on these results may move between cohorts. Any results from testing undertaken for other reasons (e.g. symptoms, contact tracing etc.) or prior to study entry will also be included. Individuals will complete enrolment and fortnightly questionnaires on exposures and symptoms. Follow-up will be for at least 12 months from study entry.

**Outcome:** The primary outcome of interest is a reinfection with SARS -CoV-2 during the study period. Secondary outcomes will include incidence and prevalence (both RNA and antibody) of SARS-CoV-2, viral genomics, viral culture, symptom history and antibody/neutralising antibody titres.

**Conclusion:** This large study will help us to understand the impact of the presence of antibodies on the risk of reinfection with SARS-CoV-2; the results will have substantial implications in terms of national and international policy, as well as for risk management of contacts of COVID-19 cases.

**Trial Registration:** IRAS ID 284460, HRA and Health and Care Research Wales approval granted 22 May 2020.

## 1. Introduction

SARS-CoV-2, a novel coronavirus which causes respiratory illness, was first identified in China in December 2019.(1) Following global spread of the virus, the World Health Organization declared a national pandemic in March 2020. Globally nearly 64 million cases have been reported to the World Health Organization by 4 December 2020, with 1,488,120 deaths attributed to COVID-19 (2), and both the virus and the measures put in place to reduce spread have led to significant economic and societal impacts. Whether individuals can be re-infected with SARS-CoV-2 is a crucial question both for contact management of individuals exposed to the virus, but also from the perspective of the implications for the effectiveness of any vaccine produced.

The risk of reinfection for individuals who have previously had COVID-19 is unknown. There have been a number of case reports which have identified individuals who have been reinfected with a new and genetically distinct SARS-CoV-2 genome from their original infection.(3-8) One recent study of the antibody prevalence from three large cross-sectional surveys in England, measured using a self-administered test, found that over a three month period population prevalence dropped from 6.0% (5.8, 6.1) to 4.4% (4.3, 4.5), suggesting waning antibodies in the population.(9) Several longitudinal studies have looked at titres over time, with one UK based study showing waning of neutralising antibodies over 3 months but with large differences between individuals (those with more severe disease had higher antibody titres at their peak)(10), while another study in Iceland demonstrated maintenance of IgG titres over 4 months.(11) However the implications of these findings are unclear. We know that other human coronaviruses demonstrate similar patterns of waning titres over time, with individuals able to be reinfected and shed virus.(12)

Many hospitals are choosing to screen certain staff groups regularly for SARS-CoV-2 to reduce the risk of transmission to patients and colleagues. Healthcare workers have consistently been found to have higher positive antibody prevalence compared with the general population; published surveys in UK hospital staff have reported prevalences of 24.5% in a Birmingham Hospital(13), and 31%(14) and 44%(15) in London. For a study examining the risk of reinfection an ideal population to examine this question is one with a high baseline antibody prevalence, where there is an indication for routine SARS-CoV-2 screening, easy access to testing and likely to be higher ongoing exposure to SARS-CoV-2 in hospitals.

The SIREN (**S**arscov2 **I**mmunity & **RE**infection Evaluatio**N**) study aims to answer the key question of whether prior SARS-CoV-2 infection confers future immunity to SARS-CoV-2 reinfection. The study design will also enable important secondary outcomes to be examined, including antibody titre change over time, incidence of new infections, clinical and demographic factors correlating with antibody presence, phylogenetic relatedness of healthcare worker infections and ability to culture viable virus from those who are reinfected.

## 2. Methods

### 2.1. Study design

This is a prospective longitudinal cohort study which will enrol up to 100,000 individuals and follow them up for 12 months with regular data collection. Individuals will be enrolled between June 2020 and March 2021.

### 2.2. Study objectives

The overall aim of this study is to determine if prior SARS -CoV-2 infection in health care workers confers future immunity to reinfection.

#### Primary Objective

To determine whether the presence of antibody to SARS-CoV-2 (anti-SARS-CoV-2) is associated with a reduction in the subsequent risk of reinfection over short term periods (reviewed monthly) and the next year.

#### Secondary Objectives

1. To estimate the prevalence of SARS-CoV-2 infection in staff working in healthcare organisations by region, using baseline serological testing at study entry and symptom history from January 1st 2020 to date of study entry
2. To estimate the subsequent incidence of symptomatic and asymptomatic SARS-CoV-2 infection and determine how this varies over time, using regular PCR testing (combined with any intercurrent symptomatic testing)
3. To estimate cumulative incidence of new infections in staff working in healthcare organisations stratified by age, sex, staff group, ethnicity and co-morbidities
4. To measure the ability to culture viable virus from cases of reinfection diagnosed by PCR and whether those who are persistently positive on PCR are continuing to shed viable virus
5. To use genomic comparison to determine whether healthcare workers who become PCR-positive for a second time within a defined time frame are experiencing persistent infection or reinfection
6. To determine how serological response changes over time
7. To determine whether there is a relationship between serological response (using enzyme immunoassay detection of IgG) and the presence of neutralising (protective) antibodies
8. To identify serological, demographic or clinical factors that correlate with the presence of neutralising antibodies, including subsequent disease severity
9. To investigate the phylogenetic relatedness of SARS-CoV-2 viruses causing staff working in healthcare organisations infections

### 2.3. Participants and recruitment

#### Population

The eligible population are staff members of healthcare organisations. Staff are recruited from healthcare organisations participating as SIREN sites, and all NHS Trusts/Health Boards (organisations that manage hospitals) in England, Scotland, Wales and Northern Ireland have been invited to join. At a later stage, recruitment may be extended to staff from other healthcare organisations such as primary care organisations and the independent sector.

#### Eligibility

A participants is eligible to join the study if they are a healthcare organisation staff member who works in a clinical setting where patients are present, can provide written consent, and is willing to remain engaged with follow-up for 12-months. Temporary short-term staff members are not eligible.

#### Recruitment and consent

Sites are responsible for recruiting eligible participants, according to their own processes. Sites are recommended to circulate all staff communications inviting volunteers and to monitor the demographics of their cohort as they recruit, aiming to represent their staff population. There are no requirements for quotas or structured sampling.

Interested and eligible potential participants are provided with a unique study number and passcode by their site research team and directed to enrol in the study by completing the online consent form and enrolment questionnaire. On completion of the online consent form and enrolment questionnaire, participants join the SIREN cohort. Site research teams are automatically informed of participant enrolment in real-time, and can then contact participants to arrange testing.

### 2.4. Data collection

#### At enrolment

At enrolment participants complete an online questionnaire and submit serum and a nose swab (or nose and throat swab) for SARS-CoV-2 antibody and nucleic acid amplification (NAAT) testing. Participants will have up to 10mls of blood taken by venepuncture at enrolment and follow-up. The questionnaire collects information on participant demographics, work environment, symptom and testing history, participation in clinical trials and known COVID exposures since 1 January 2020.

#### At follow-up

Participants undergo regular repeat NAAT and antibody testing throughout the study period, initially at fortnightly intervals, although frequency may be revised (weekly to monthly) subject to local/national epidemiology and feedback. Participants are sent a link to an online follow-up questionnaire on a fortnightly basis, with a reminder message sent after 2 days if the follow-up questionnaire is not completed. These questionnaires capture information on symptoms, exposures and subsequent enrolment in vaccine or prophylaxis trials.

#### Testing at SIREN site laboratories and data sources

For all participants NAAT (typically PCR) and antibody testing is undertaken locally at the laboratory used by their healthcare organisation. The healthcare organisation is responsible for issuing results to the participants as per local procedures. Testing platforms, including choice of antibody assay, is determined locally.

All laboratories for SIREN participating sites submit their antibody and antigen testing data into Public Health England’s (PHE) Second Generation Surveillance System (SGSS). Testing data from sites on SIREN participants is obtained by the PHE SIREN team through deterministic linkage, based on the NHS number (or equivalent unique identifier for Devolved Administrations) and additional patient identifiers provided by participants in the enrolment questionnaire. Linkage to site testing data for participants in Devolved Administrations is organised with the support of their respective public health agencies. At enrolment, participants consent for the SIREN team to link all their historic and future SARS -CoV-2 testing data, including tests undertaken prior to enrolment, and tests taken outside SIREN, such as tests taken due to symptoms or exposures.

### 2.5. Testing at Public Health England

#### Serology

For all participants, at enrolment an aliquot of 2ml serum will be shipped to and stored in the PHE biobank. At follow-up, serum samples for participants who have ever been antibody positive or antigen positive or have enrolled in a vaccine trial will be sent to and stored at the PHE biobank.

At enrolment, all participants will have their serum re-tested by PHE for antibodies to SARS-CoV-2, including the Roche Elecsys Anti-SARS-CoV-2 spike (S) and nucleocapsid (N) protein assays(16) and additional in-house assays to examine for neutralising antibody. Individuals will be classified as seropositive or seronegative based on PHE antibody testing for N and S.

In addition to the cohort serological characterisation at enrolment described above, seropositive participants in whom reinfection is identified, plus a cohort of matched non-infected seropositive controls, will have their sera further characterised using additional assays and for the presence of neutralising antibody, to provide hypothesis generating data on mechanisms of protective immunity.

#### Genomic analysis

All positive samples from participants will be sequenced as part of the routine sequencing of NHS residual samples in COG-UK Consortium laboratories. For participants who have more than one positive PCR test, genomes will be compared where possible to provide evidence to support reinfection or persistent infection. Phylogenetic analysis of SARS -CoV-2 from staff in healthcare organisations, using the study samples and the wider collection of genomes available through the COG-UK Consortium, will also be undertaken as an exploratory analysis into the diversity and spread of SARS -CoV-2 in healthcare workers.

#### Viral Culture

Participants with possible reinfection or persistent infection will be identified and viral culture requested. This may be on residual sample from the swab already taken, but in certain circumstances (e.g. viral culture not possible on the residual sample) we may request another swab is taken and sample sent for culture.

#### T-cell assays and other studies

Participants who are persistently NAAT positive, have potentially been reinfected, or have discordant serology may be contacted by the SIREN Study Team to link into optional regional sub-studies e.g. assessing T cell assays and antibody dynamics.

### 2.6. Sample size and power

A simulation approach using a mixed effects Poisson regression model has been used to estimate the power to detect relative differences between the study cohorts. Our key assumptions include that 25% of our cohort will be seropositive at enrolment (based on 20% of staff who were asymptomatic and tested positive in one London hospital between 23 March and 2 May 2020(14)), and a total attrition of 35%, (unaffected by serostatus and occurring at a constant rate). The proportion of seropositive recruits at each site has been obtained from a Gaussian distribution with a mean of 0.25 and standard deviation of 0.05 to reflect expected inter-site variation.

Power was estimated as the proportion of simulations for which the Wald statistic p value for the estimated incidence rate ratio in the seropositive compared to seronegative cohorts was less than 0.05. Our simulations found that there is statistical power of 80% or greater to detect a relative decrease of 30% or greater in cumulative incidence, provided the cumulative incidence in the seronegative group is in excess of 5%; even taking the cumulative incidence to as low as 2% in the seronegative group there is still sufficient power of in excess 80% for relative decrease of 80% or greater.

It was assumed that on average 250 participants would be recruited from each selected healthcare organisation, with a standard deviation of 50. The cumulative incidence in each site in the seronegative cohort has been simulated using Gaussian distributions with means of 0.05, 0.1, 0.2 and 0.3 each with a coefficient of variation of 0.2. This range represents that which is feasible to observe over a 12-month period, given the behavioural and social interventions still being employed during the study to control transmission.

A study duration of 52 weeks has been assumed with the inter-test period of 2 weeks.

It was assumed that the cumulative incidence in the seronegative cohort was 30% with a between trust coefficient of variation of 0.1, reflecting levels of seropositivity in HCWs at the time. Relative reductions in cumulative incidence in the seropositive cohort was varied between 1 (no protection from infection) to 0.1 (antibody effectiveness of 90%). Units in the simulations were allocated to be infected or not, using a draw from a Bernoulli distribution with p equal to the site and cohort specific simulated cumulative infection rate. A simplifying assumption of a constant infection rate over the study period has been used.

For each scenario a set of 200 simulations were performed. For each simulation, the total number if infections and person weeks of follow-up was calculated for each cohort in each organisation. This data was analysed using a mixed effects Poisson model, using the natural logarithm of the person weeks as an offset. These are presented in Table 1, indicating that there is sufficient power for all but the smallest immune efficacy of 0.1 i.e. a 10% reduction in incidence in the seropositive cohort. Such a small reduction is indicative of a level of protection unable to provide a means of controlling the pandemic via natural herd immunity.

**Table 1:** Power estimates obtained via simulation for a range of immune effectiveness and cumulative incidence.

| Cumulative incidence in the seronegative at baseline cohort (per 100 participants) in 12 months | Immune Effectiveness |  |  |  |  |
| --- | --- | --- | --- | --- | --- |
|  | 10% | 20% | 30% | 40% | 50% |
| 0.05 | 0.15 | 0.44 | 0.79 | 0.98 | 1.00 |
| 0.1 | 0.20 | 0.77 | 0.99 | 1.00 | 1.00 |
| 0.2 | 0.53 | 0.99 | 1.00 | 1.00 | 1.00 |
| 0.3 | 0.67 | 1.00 | 1.00 | 1.00 | 1.00 |

### 2.7. Statistical Analysis Plan: primary outcome measure

All enrolled participants will be included in analyses, which will account for clustering by research site. Analyses will be conducted at regular intervals following sufficient events of interest.

Estimates of both cumulative incidence and incidence density in the seropositive and seronegative cohorts will be obtained using mixed effects models assuming counts of PCR positive have a negative binomial distribution, a log link function, and the natural logarithm of the total number of subjects or the total follow-up time use as an offset, respectively. Inclusion of a binary predictor indicating the serostatus of the cohort into this model will provide estimates of the incidence rate ratio. Sites will be incorporated as a random intercept to account for unmeasured, shared, site level factors. To account for a non-constant force of infection, calendar month will be incorporated as an additional random effect. An assessment of the role of factors such as age, gender and ethnicity in immunity will be explored by inclusion of interactions within the model between each and serological status.

While the above analytical approaches provide a “classical” person-years approach to prospective cohort analysis and provide familiar measures of association, it may be inadequate to assessment of immunity provided by seroconversion. As it is expected that seropositivity is likely to confer a degree of short to median term protection for a SARS-CoV-2 infection, multi-state and parametric cure rate models incorporating frailty will also be employed. Bayesian approaches to cure rate models with frailty as describe by deSouza(17) will be employed.

It is also possible to introduce “misclassification” of state into the multi state model, providing an estimate of sensitivity to account for imperfect serological tests. Approaches like those proposed by Jackson(18) will be employed.

#### Procedure for Accounting for Missing, Unused, and Spurious Data

Analyses will be restricted to cases with antibody and PCR tests. The PCR test for virus is being used as a diagnostic test and hence has high performance. Sufficient sera will be obtained to re-run the immunological assays in case of initial assay failure. For similar reasons we do not anticipate that spurious data will be obtained.

#### Procedures for Reporting any Deviation(s) from the Original Statistical Plan

Deviations from the original statistical plan or the statistical analysis plan will be described and justified in the analysis reports.

Data will be analysed using STATA.v15 and R software.

## 3. Study oversight

Oversight is provided by the Study Management Group, chaired by the Chief Investigator, with representatives from Public Health England, Public Health Scotland, Public Health Wales, Public Health Agency (Northern Ireland), and the COVID-19 Genomics Consortium UK (COG-UK).

The study follow-up period will end by default 12 months following the enrolment of the last participant, but by consensus of the Study Management Group and funder may be terminated sooner if findings are sufficient. There are no formal stopping rules for futility, utility or lack of power. The final decision to terminate the study will be made by Public Health England and Department for Health and Social Care.

## 4. Ethics and Consent

The study has received approval from Berkshire Research Ethics Committee and has also received support from NIHR as an urgent public health study, which allows central research network resources to recruit participants. All participants have provided informed consent prior to entry to the study and have the option to withdraw at any time. At withdrawal, participants can choose to have their data or samples retained or destroyed, or partial variations. Protocol deviations and breaches will be recorded by the site research teams and the Sponsor will be informed of any serious breaches within one working day.

## 5. Discussion

### 5.1. Strengths

This study is the largest national longitudinal study of this scale examining the question of reinfection with SARS -CoV-2 that the authors are aware of globally. In a system where staff members may be tested in different settings depending on the timing and reasons for testing (community testing hubs, other hospitals, primary care), the automated method of data extraction and access to national testing data means that the study is less likely to miss potential cases. As far as possible the study is designed to run alongside normal laboratory processes; laboratories use the same assays and procedures which are in place for all other testing, reducing additional burden on sites.

The study design lends itself to forming sub-cohorts for more detailed investigations. It has active research collaborations with immunology researchers from the UK Research and Innovation (UKRI) Immunology consortium to investigate T cell responses and with the Wellcome Trust funded Humoral Immune Correlates of COVID-19 (HICC) consortium to investigate humoral immune responses.

### 5.2. Weaknesses

Cohort retention will be an important consideration for the study team, to avoid losing power to detect the primary outcome and potential introduction of bias if there is differential attrition by cohort. To mitigate this, the study team will actively monitor withdrawals and participant feedback, to implement improvements and will establish direct participant communications (e.g. a newsletter) to promote engagement. Over the study period, it is likely that vaccine trials and usage will increase; adjustments to the study methodology may be required to permit co-enrolment and retain SIREN participants who subsequently receive vaccines, and to incorporate vaccine efficacy into the analyses. Differences in demographics, general health and ongoing risk of exposure between healthcare workers and the general population mean that the results may not be fully generalisable to the UK population.

## Data Availability

Not applicable

## 6. Declarations

### 6.1. Ethics

The study has received ethical approval from Berkshire Research Ethics Committee (20/SC/0230). Study participants will provide informed written consent prior to study entry.

### 6.4. Authors’ contributions

SH is the Chief Investigator and conceived the study. SH, CB, and MAC designed the study and drafted the first protocol, with substantial design contribution from MR, MZ and TB. AC wrote the statistics plan and power calculations. VH, SW, MJC, PK, MS, SR, BO and AV were responsible for designing or updating aspects of the study design. SW and VH re-drafted the most recent protocol. SH, SW and VH prepared the manuscript for publication, with the review and approval of the other authors.

#### 6.5. Authors’ Acknowledgements

The authors would like to thank our partners in the Devolved Administrations, particularly Lesley Price and Muhammad Sartaj, for their contribution and advice.

### 6.6. Availability of Data

Not applicable

### 6.7. Consent for Publication

Not applicable

## References

1. Government of the Hong Kong Special Administrative Region. CHP closely monitors cluster of pneumonia cases on Mainland 31/12/2019 2019 [updated 31/12/2019]. Available from: https://www.info.gov.hk/gia/general/201912/31/P2019123100667.htm.

2. World Health Organization. COVID-19 Dashboard 2020 [4/12/2020]. Available from: https://covid19.who.int/.

3. Van Elslande J, Vermeersch P, Vandervoort K, Wawina-Bokalanga T, Vanmechelen B, Wollants E, et al. Symptomatic SARS-CoV-2 reinfection by a phylogenetically distinct strain. Clin Infect Dis. 2020.

4. To KK, Hung IF, Ip JD, Chu AW, Chan WM, Tam AR, et al. COVID-19 re-infection by a phylogenetically distinct SARS-coronavirus-2 strain confirmed by whole genome sequencing. Clin Infect Dis. 2020.

5. Prado-Vivar B, Becerra-Wong M, Guadalupe JJ, Márquez S, Gutierrez B, Rojas-Silva P, et al. A case of SARS-CoV-2 reinfection in Ecuador. Lancet Infect Dis. 2020.

6. Larson D, Brodniak SL, Voegtly LJ, Cer RZ, Glang LA, Malagon FJ, et al. A Case of Early Re-infection with SARS-CoV-2. Clin Infect Dis. 2020.

7. Tillett RL, Sevinsky JR, Hartley PD, Kerwin H, Crawford N, Gorzalski A, et al. Genomic evidence for reinfection with SARS-CoV-2: a case study. Lancet Infect Dis. 2020.

8. Gupta V, Bhoyar RC, Jain A, Srivastava S, Upadhayay R, Imran M, et al. Asymptomatic reinfection in two healthcare workers from India with genetically distinct SARS-CoV-2. Clin Infect Dis. 2020.

9. Ward H, Cooke G, Atchison C, Whitaker M, Elliott J, Moshe M, et al. Declining prevalence of antibody positivity to SARS-CoV-2: a community study of 365,000 adults. medRxiv.yv2020:2020.10.26.20219725.

10. Seow J, Graham C, Merrick B, Acors S, Pickering S, Steel KJA, et al. Longitudinal observation and decline of neutralizing antibody responses in the three months following SARS-CoV-2 infection in humans. Nat Microbiol. 2020;5(12):1598–607.

11. Gudbjartsson DF, Norddahl GL, Melsted P, Gunnarsdottir K, Holm H, Eythorsson E, et al. Humoral Immune Response to SARS-CoV-2 in Iceland. N Engl J Med. 2020;383(18):1724–34.

12. Sariol A, Perlman S. Lessons for COVID-19 Immunity from Other Coronavirus Infections. Immunity. 2020;53(2):248–63.

13. Shields A, Faustini SE, Perez-Toledo M, Jossi S, Aldera E, Allen JD, et al. SARS-CoV-2 seroprevalence and asymptomatic viral carriage in healthcare workers: a cross-sectional study. Thorax. 2020;75(12):1089–94.

14. Grant JJ, Wilmore SMS, McCann NS, Donnelly O, Lai RWL, Kinsella MJ, et al. Seroprevalence of SARS-CoV-2 antibodies in healthcare workers at a London NHS Trust. Infect Control Hosp Epidemiol. 2020:1–3.

15. Houlihan CF, Vora N, Byrne T, Lewer D, Kelly G, Heaney J, et al. Pandemic peak SARS-CoV-2 infection and seroconversion rates in London frontline health-care workers. Lancet. 2020;396(10246):e6–e7.

16. Public Health England, University of Oxford. Evaluation of sensitivity and specificity of four commercially available SARS-CoV-2 antibody immunoassays. 2020.

17. de Souza D, Cancho VG, Rodrigues J, Balakrishnan N. Bayesian cure rate models induced by frailty in survival analysis. Stat Methods Med Res. 2017;26(5):2011–28.

18. Jackson CH. Multi-State Models for Panel Data: The msm Package for R. Journal of Statistical Software. 2011;38(8):1–28.

